# A dynamic model to estimate evolving risk of major bleeding after percutaneous coronary intervention

**DOI:** 10.1101/2021.12.17.21267935

**Authors:** Nathan C Hurley, Nihar Desai, Sanket S. Dhruva, Rohan Khera, Wade Schulz, Chenxi Huang, Jeptha Curtis, Frederick Masoudi, John Rumsfeld, Sahand Negahban, Harlan M. Krumholz, Bobak J. Mortazavi

## Abstract

**Background:** Bleeding is a complication of percutaneous coronary intervention (PCI), leading to significant morbidity, mortality, and cost. Existing risk models produce a single estimate of bleeding risk anchored at a single point in time and do not update estimates as clinical information emerges, despite the dynamic nature of risk.

**Objective:** We sought to develop models that update estimates of bleeding risk over time, incorporating evolving clinical information, and to demonstrate updated predictive performance.

**Methods:** Using data available from the National Cardiovascular Data Registry (NCDR) CathPCI, we trained 6 different tree-based machine learning models to estimate the risk of bleeding at key decision points: 1) choice of access site, 2) prescription of medication prior to PCI, and 3) the choice of closure device.

**Results:** We included 2,868,808 PCIs; 2,314,446 (80.7%) prior to 2014 for training and 554,362 (19.3%) remaining for validation. Discrimination improved from an AUROC of 0.812 (95% Confidence Interval: 0.812-0.812) using only presentation variables to 0.845 (0.845-0.845) using all variables. Among 123,712 patients classified as low risk by the initial model, 14,441 were reclassified as moderate risk (1.4% experienced bleeds), while 723 were reclassified as high risk (12.5% experienced bleeds). Among 160,165 patients classified as high risk by the initial model, 40 were reclassified to low risk (0% experienced bleeds), and 43,265 to moderate risk (2.5% experienced bleeds).

**Conclusion:** Accounting for the time-varying nature of data and capturing the association between treatment decisions and changes in risk provide up-to-date information that may guide individualized care throughout a hospitalization.

**Condensed Abstract:** Existing risk models for bleeding with PCI produce a single estimate anchored at a single point in time. We developed models that update estimates of bleeding risk over time, incorporating evolving clinical information, using data available from the National Cardiovascular Data Registry (NCDR) CathPCI. We trained 6 different machine learning models to estimate the risk of bleeding at key decision points, improving discrimination from an AUROC of 0.812 to 0.845, over time. Accounting for the time-varying nature of data and capturing association between treatments and changes in risk provide up-to-date information that may guide individualized care throughout a hospitalization.

## Introduction

Bleeding is a common complication of percutaneous coronary intervention (PCI), leading to significant morbidity, mortality, and cost (1). Several tools have been developed to predict post-PCI bleeding, including two models (2-4) from the American College of Cardiology’s National Cardiovascular Data Registry (NCDR) (5). By informing clinicians about bleeding risk, these models can aid the selection of bleeding avoidance strategies, such as combination of radial access and the use of bivalirudin interprocedurally as well as post-procedural care strategies, particularly in high-risk patients, thereby reducing rates of major bleeding complications and improving care quality and clinical outcomes (6,7). A problem with existing models is that they produce a single estimate of bleeding risk anchored at a single point in time, i.e., data prior to the procedure inform a single estimate for risk of post-PCI bleeding. These models do not update the risk estimates as new clinical information emerges. Therefore, as treatment decisions are made or unforeseen events occur, these models are unable to adapt and incorporate new information.

Risks are dynamic in nature; as data are gathered and treatment decisions are made, risk estimates should account for all the information currently available, including changes in clinical status of patients as a response to treatment decisions (8). Dynamic models can enable estimation of risk that adapts and updates throughout an episode of care. For example, in the ACUITY trial that evaluated the use of bivalirudin in ACS, intra-procedural events were strongly associated with adverse outcomes (9), which would be expected to alter antiplatelet and anticoagulant strategies. We hypothesize that bleeding risk is similarly a dynamic process, affected by multiple pre-, intra-, and post-PCI patient and procedural factors throughout the care pathway.

Risk models that update across the patient episode of care have the potential to improve our ability to individualize risk prediction. Providing physicians up-to-date feedback may inform optimization of therapeutic strategies through enhanced decision-support at actionable points during an episode of care involving a PCI procedure. These models may also improve the understanding of the dynamics and key variables affecting bleeding risk, representing a transformational change in risk prediction and embrace the principles of a learning health care system (10). Accordingly, we sought to develop models that update estimates of patient risk of bleeding over time, enabling a dynamic estimate of risk that incorporates evolving clinical information.

## Methods

### Study cohort

This study included all index PCIs in the National Cardiovascular Data Registry (NCDR) CathPCI registry version 4.4 from July 2009 through April 2015 (5,11). To examine the improvement of dynamic data over static risk prediction models, we used the same cohort as Mortazavi et al. (4). Therefore, we excluded patients during readmissions, who died in the hospital, who had missing data regarding if they had any bleeding events, or who underwent coronary artery bypass grafting (CABG) during the index admission (3,4).

The primary outcome was any in-hospital bleeding event within 72 hours after the start of the PCI procedure. Bleeding was defined using the definition employed in prior NCDR risk models as a hemoglobin drop ≥3 g/dL, whole blood or packed red blood cell transfusion, or intervention/surgery at the bleeding site to reverse/stop or correct bleeding. We further excluded patients with multiple, unknown, or brachial access sites to evaluate the treatment decision point of radial versus femoral access. We additionally excluded patients with multiple closure methods. This final exclusion was added because multiple closure methods may be associated with multiple access or may represent bleeding (e.g., a femoral closure device did not work and so manual pressure was held), which would bias the model to predicting risk of bleeding after the bleed has occurred.

### Variables of Interest

This study considered all data available from the CathPCI Registry prior to patient discharge (5). This included all data used by the existing models (3,4), as well as additional variables as described below. The current full existing NCDR bleeding risk model (3) uses 31 variables: 23 patient characteristics at the time of presentation and 8 characteristics related to coronary anatomy and lesion characterization (see **Supplementary Material**). A more recent work added additional variables from the registry related to those 31 variables, finding improved predictive performance (4). Our study includes further additional data. The additional data considered in this study consists of additional laboratory data, past medical history, additional coronary anatomy (including percent stenosis), stent type, and closure method categories (manual compression, sealant, mechanical, suture, patch, staple, other, or none).

### Staged Model Analysis

We sorted all available CathPCI data into key decision stages of a PCI episode of care. First, we defined three decision points that affect bleeding risk: 1) choice of access site (radial versus femoral); 2) choice of medications (including those administered 24 hours pre-procedure and intra-procedure); and 3) choice of closure device indicator variables. While the choice of closure for radial access would be expected to be none, there are closure devices used with Radial Access. Without an ability to do a chart review, we take this data as is (and discuss further in the **Supplementary Material**).

Using these three key decision points, we evaluated variables available at three stages: 1) variables available at patient presentation prior to PCI; 2) variables available after diagnostic coronary angiography; and 3) variables related to the PCI procedure. Combining these three decision points and the information available at each of them, six models were designed (**Figure 1**). The first included only variables available at presentation to the cardiac catheterization lab. Each subsequent model added either a decision node or information that could inform the next decision. The final model included all variables and clinical decisions through the PCI procedure, evaluating remaining bleeding risk for post-procedural care.

**Figure 1.**
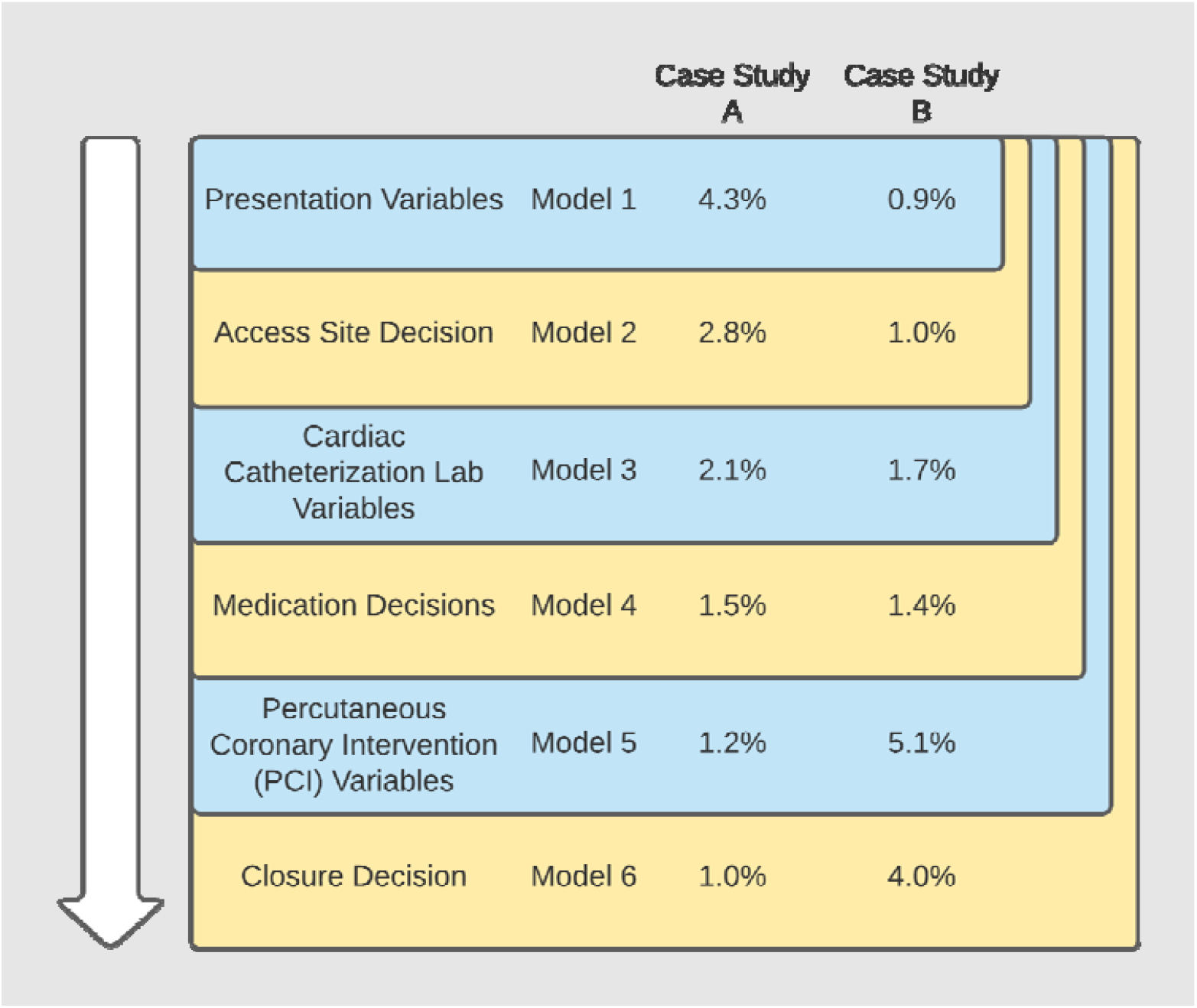
Model hierarchy. Each model integrated information of all features from prior models, as well as an added set of features. Passage through an episode of care proceeds from the top to the bottom. Case study risks over time at each stage are shown here.

### Data Preparation

The NCDR utilizes a high-quality review and adjudication process, ensuring minimal missing data across variables (12). Additionally, several steps in data preparation were necessary prior to model development, including data cleaning to determine the true rate of missing, including for daughter variables only collected if the indicator variable suggests it should have. Details are provided in the **Supplementary Material**.

Once data was cleaned, the missing values were imputed using multiple multivariate feature imputation. Each missing feature was modeled using Bayesian ridge regressors trained in a round-robin fashion. This imputation was performed using the Iterative Imputer package in scikit-learn 0.24.1 (13), based on the multivariate imputation by chained equations (mice) package for R (14). Following imputation, binary and ordinal variables were set to the nearest allowed value. Multiple imputations were produced by sampling from the regressor models multiple times; each discrete sampling was a new overall sample from the model. This sampling was used to produce five folds of imputations. We used a re-imputation technique for handling the longitudinal nature of the data, producing multiple imputed datasets at each stage through an episode of care (15). The imputation models were trained on the training dataset prior to cross validation (16). The test sets were multiply imputed, but the regressors used for this imputation were trained only on training data to avoid data leakage.

### Training, Testing, and Evaluating

The cohort was divided into temporal subsets: an initial 80% subset (July 1, 2009 through December 31, 2013) for model training and a later 20% subset (January 1, 2014 through December 31, 2014) for model validation (17,18). This temporal split allows for the fairest possible evaluation of the model through testing it on the most distinct subset. This approach protects against biasing the result of the overall model by including more recent data, as in other recent NCDR models (19). While an external validation set would be preferable, this method of separating data allows for the most realistic testing of the model (testing models trained on retrospective data on newly-seen patients). Patient characteristics for both the entire dataset as well as the training and validation splits are shown in **Table 1**.

**Table 1.** Patient characteristics.

|  | Overall<br>(n=2,868,808) | Training<br>(n=2,314,446) | Validation<br>(n=554,362) |
| --- | --- | --- | --- |
| **Demographics** |  |  |  |
| Age, mean (SD), y | 64.6 (12.0) | 64.6 (12.0) | 64.9 (11.9) |
| Men | 1,960,409 (68.3%) | 1,577,369 (68.2%) | 383,040 (69.1%) |
| BMI, mean (SD) | 30.0 (6.4) | 30.0 (6.4) | 30.1 (6.4) |
| **Cardiovascular Comorbidities** |  |  |  |
| Diabetes | 1,057,221 (36.9%) | 844,928 (36.5%) | 212,291 (38.3%) |
| Hypertension | 2,353,798 (82.1%) | 1,895,949 (81.9%) | 457,849 (82.6%) |
| Peripheral Vascular Disease | 339,316 (11.8%) | 274,039 (11.8%) | 65,278 (11.8%) |
| Chronic Kidney Disease | 861,391 (30.0%) | 705,765 (30.5%) | 155,626 (28.1%) |
| Previous PCI | 1,178,346 (41.1%) | 948,367 (41.0%) | 229,978 (41.5%) |
| Previous CABG | 510,781 (17.8%) | 414,560 (17.9%) | 96,222 (17.4%) |
| **PCI Procedural Status** |  |  |  |
| Elective | 1,196,485 (41.7%) | 992,525 (42.9%) | 203,961 (36.8%) |
| Urgent | 1,152,328 (40.2%) | 906,226 (39.2%) | 246,101 (44.4%) |
| Emergent | 512,404 (17.9%) | 409,659 (17.7%) | 102,744 (18.5%) |
| Salvage | 6,440 (0.2%) | 5,029 (0.2%) | 1,411 (0.3%) |
| Unknown | 1,150 (0.04%) | 1,007 (0.04%) | 145 (0.03%) |
| STEMI | 468,270 (16.3%) | 373,792 (16.2%) | 94,477 (17.0%) |
| Cardiogenic Shock | 64,743 (2.3%) | 51,689 (2.2%) | 13,055 (2.4%) |
| Cardiac arrest within 24h of PCI | 49,008 (1.7%) | 38,840 (1.7%) | 10,168 (1.8%) |
| Preprocedural hemoglobin, median (IQR), g/dL | 13.7 (12.4-14.9) | 13.7 (12.4-14.9) | 13.7 (12.4-14.9) |
| **Access Site** |  |  |  |
| Femoral | 2,394,173 (83.5%) | 1,997,049 (86.3%) | 397,124 (71.6%) |
| Radial | 474,635 (16.5%) | 317,397 (13.7%) | 157,238 (28.4%) |
| **Medications Used** |  |  |  |
| Ticlopidine | 5,895 (0.2%) | 5,186 (0.2%) | 709 (0.1%) |
| Clopidogrel | 1,988,178 (69.3%) | 1,654,031 (71.5%) | 334,147 (60.3%) |
| Prasugrel | 433,079 (15.1%) | 339,902 (14.7%) | 93,177 (16.8%) |
| Ticagrelor | 167,838 (5.9%) | 80,318 (3.5%) | 87,520 (15.8%) |
| Fondaparinux | 15,816 (0.6%) | 14,837 (0.6%) | 979 (0.2%) |
| Low Molecular Weight Heparin | 272,261 (9.5%) | 224,180 (9.7%) | 48,081 (8.7%) |
| Unfractionated Heparin | 1,528,882 (53.3%) | 1,197,304 (51.7%) | 331,578 (59.8%) |
| Bivalirudin | 1,695,225 (59.1%) | 1,375,031 (59.4%) | 320,194 (57.8%) |
| GP IIb/IIIa | 677,865 (23.6%) | 576,753 (24.9%) | 101,112 (18.2%) |
| Direct Thrombin Inhibitor | 29,512 (1.0%) | 24,961 (1.1%) | 4,551 (0.8%) |
| Closure Method |  |  |  |
| Manual Compression | 965,618 (33.7%) | 815,354 (35.2%) | 150,264 (27.1%) |
| Sealant | 916,374 (31.9%) | 745,456 (32.2%) | 170,918 (30.8%) |
| Mechanical | 512,968 (17.9%) | 358,927 (15.5%) | 154,041 (27.8%) |
| Suture | 264,494 (9.2%) | 215,420 (9.3%) | 49,074 (8.9%) |
| Patch | 99,690 (3.5%) | 84,853 (3.7%) | 14,837 (2.7%) |
| Staple | 184 (0.0%) | 171 (0.0%) | 13 (0.0%) |
| Other | 94,818 (3.3%) | 82,626 (3.6%) | 12,192 (2.2%) |
| None/Missing | 14,662 (0.5%) | 11,639 (0.5%) | 3,023 (0.5%) |
| Post-PCI Major Bleeds | 118,327 (4.1%) | 98,167 (4.2%) | 20,160 (3.6%) |

As discussed above, five imputation folds were created for each stage. The model used was XGBoost, a gradient descent boosted decision tree model (20). We conducted an internal five-fold cross-validation to tune the hyperparameters of the model for each stage and each fold. The hyperparameters tuned were maximum depth of each tree (2, 4, 6, or 8), number of tree estimators (100, 500, 1000, or 5000), and learning rate for the model (0.1, 0.15, 0.2, or 0.3). This internal cross-validation was performed using a halving grid search as implemented by the scikit-learn package HalvingGridSearchCV (13). Optimal performance was found in 27 of the 30 experiments (6 stages * 5 folds) to be given by 1000 estimators with a max depth of 2 and a learning rate of 0.1.

The five imputation folds allow for training and validating the model multiple times, providing both estimates of overall model performance and uncertainty (17). From this, we calculated the area under the receiver operating characteristics curve (AUROC) for evaluating model discrimination (c-statistic). To better understand positive predictive value across the full range of risk stratification, we also calculated the area under the precision recall curve (AUPRC). Briefly, the precision-recall curve calculates the tradeoff between precision (positive predictive value) and recall (sensitivity) across the full range of thresholds (21). This model calibration also allows calculation of the Brier Skill Score, which provides an assessment of model calibration on an easy to interpret scale of 0-100% for calibration fit, as well as the Brier Decomposition (Reliability and Resolution), which provides measures of over and under estimation of the calibration curve.

### Variable Importance

Beyond model performance, model interpretation is a key factor in clinical utility (17). One approach for interpreting models is SHapley Additive exPlanations (SHAP) (22). SHAP attributes an importance value to each feature of a given set, therefore allowing for an ordering of features from greatest to least impact on model output. SHAP values were generated to provide visual understanding about the impact of factors driving changes in accuracy of the risk prediction model and decisions through the stages outlined in Figure 1.

To further understand dynamic risk predictions following a decision, shift tables were generated. For these, patients were classified into categories of low risk (<1%), moderate risk (1%-4%), or high risk (>4%) of bleeding. These thresholds were chosen to approximately balance patients between categories across all models (19). These shift tables are useful for visualizing changing risks of bleeding before and after a decision or for comparing the performance of the initial and final models.

### Case Studies

To further understand dynamic risk and changes in variable performance, we then cherry picked two example cases that had changes in risk and explain their journey through the episode of care and the results of which are described in the **Supplementary Material**.

All analyses were conducted in Python version 3.8.6 or R version 4.0.3. Data analysis was performed using scikit-learn 0.24.1 (13) and XGBoost 1.3.3 (20) for gradient descent boosting. SHAP explanations were generated and visualized with SHAP 0.38.1 (22). Model calibrations generated in R with mgcv 1.8-33 (23) and calibration variances with sandwich 3.0-0 (24). Source code is available online at: https://github.com/stmilab/DynamicBleeding/. Ethics oversight for data and analyses were provided and approved by the Yale Human Research Protection Program (IRB #0607001639).

## Results

### Patient Cohort, Variables Used, and Overall Performance

We included 2,868,808 PCIs in the NCDR CathPCI registry; 2,314,446 (80.7%) prior to 2014 for model training and 554,362 (19.3%) after 2014 for validation and model interpretation. The mean (SD) age of patients was 64.6 (12.0) years and 68.3% were male (**Table 1**). Overall, there were 118,327 (4.1%) major bleeding events: 98,167 (4.2%) major bleeding events in the training set and 20,160 (3.6%) in the validation set. Model performance metrics at each stage are provided in **Table 2**, and each stage is described further below.

**Table 2.** Comparison of model performances for bleeding prediction. We compare model performance by area under the receiver operating characteristic curve (AUROC), the area under the precision recall curve (AUPRC), the Brier Skill Score, and the Reliability and Resolution of the Brier Decomposition.

| Model | AUROC | AUPRC | Brier Skill Score | Brier Reliability | Brier Resolution |
| --- | --- | --- | --- | --- | --- |
| 1 | 0.812 (0.812-0.812) | 0.203 (0.203-0.203) | 0.088 (0.088-0.088) | 2.6E-4 (2.6E-4-2.6E-4) | 3.3E-3 (3.3E-3-3.3E-3) |
| 2 | 0.817 (0.817-0.817) | 0.204 (0.204-0.205) | 0.091 (0.091-0.091) | 2.0E-4 (1.9E-4-2.0E-4) | 3.4E-3 (3.4E-3-3.4E-3) |
| 3 | 0.825 (0.825-0.825) | 0.208 (0.208-0.208) | 0.094 (0.094-0.094) | 2.1E-4 (2.0E-4-2.1E-4) | 3.5E-3 (3.5E-3-3.5E-3) |
| 4 | 0.832 (0.832-0.832) | 0.217 (0.216-0.217) | 0.102 (0.102-0.102) | 1.4E-4 (1.3E-4-1.4E-4) | 3.7E-3 (3.7E-3-3.7E-3) |
| 5 | 0.844 (0.844-0.845) | 0.241 (0.240-0.241) | 0.118 (0.118-0.118) | 1.1E-4 (1.1E-4-1.2E-4) | 4.2E-3 (4.2E-3-4.2E-3) |
| 6 | 0.845 (0.845-0.845) | 0.242 (0.241-0.242) | 0.119 (0.119-0.119) | 1.0E-4 (1.0E-4-1.1E-4) | 4.3E-3 (4.3E-3-4.3E-3) |

### Interpreting SHAP plots

SHAP feature importance plots are shown in Figures 2-7. In each plot, the variables are sorted in order of decreasing importance. The color of a variable relates to the value of that variable, while location along the x-axis represents how much that variable contributes to the risk of bleeding. Features most strongly driving predictions of high bleeding risk appear on the right with high SHAP values, while features most predictive of low bleeding risk appear at the left. The specific plots, per stage, are described further below.

**Figure 2.**
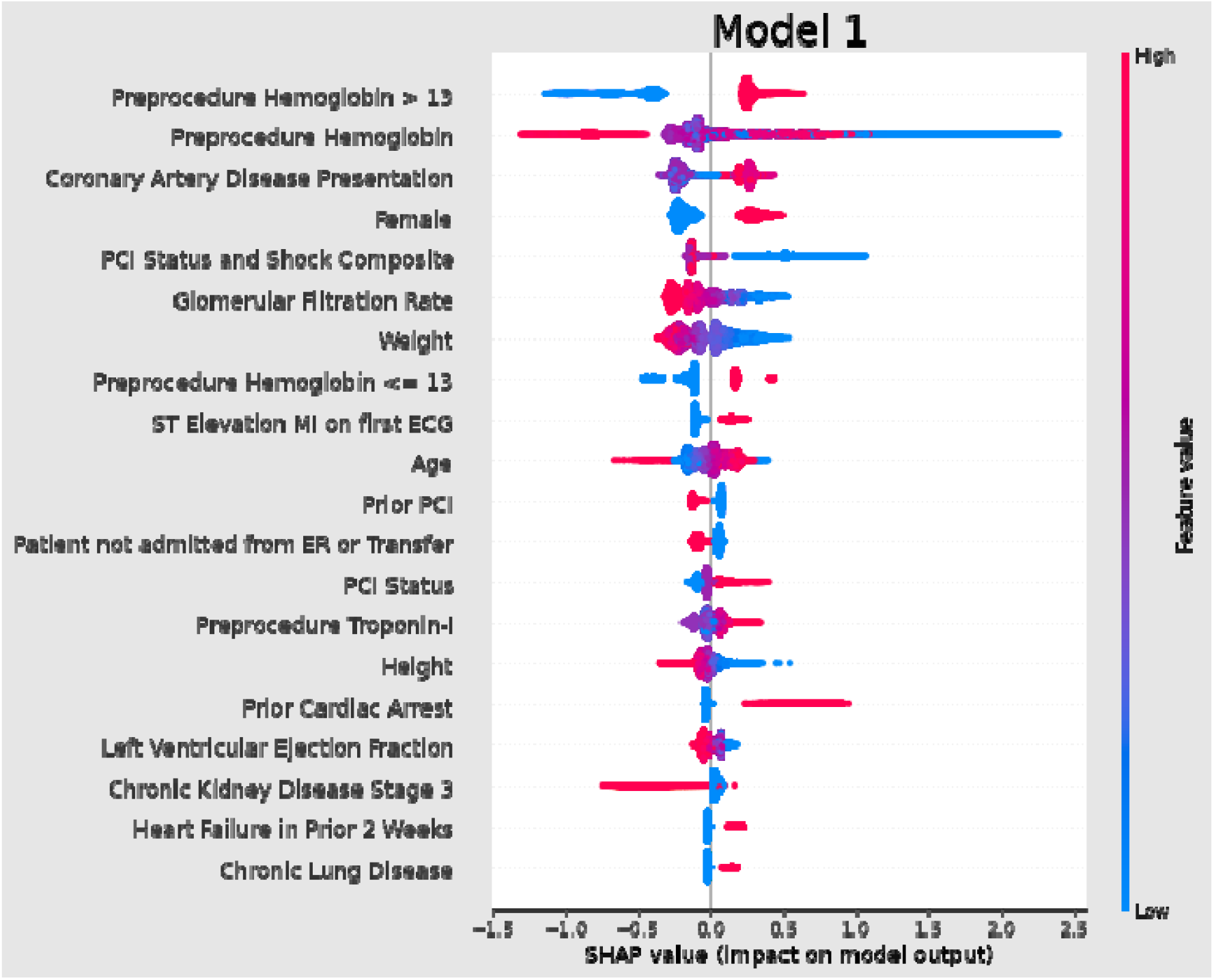
SHAP Tree explainer for Model 1. Variables are sorted in order of decreasing importance. Variable color relates to the value of that variable, while location along the x-axis represents how much that variable contributes to the risk of bleeding. Features most strongly driving predictions of high bleeding risk appear on the right, while features most predictive of low bleeding risk appear at the left. The higher a variable is, the greater overall importance that variable exhibits for the model. Red values indicate high values for that variable (if continuous or ordinal) or “true” (if binary), while blue values indicate the opposite. Points to the right of the axis (positive SHAP values) indicate that a feature of that value increases model estimate of bleeding risk, while points to the left of the axis (negative SHAP values) indicate that a feature of that value decreases the model estimate of bleeding risk. For instance, on the top row of Figure 2, a preprocedural hemoglobin greater than 13 is associated with an increased risk of bleeding, while value less than 13 is associated with a decreased risk. The wide range of SHAP values for this variable shows that while the direction of association is constant, the degree to which this feature impacts risk is not constant.

### Stage 1: Clinical Presentation (Model 1)

The initial model uses information available to a clinician at the time that a patient presents and predicted bleeding risk with an AUROC of 0.812 and AUPRC of 0.203. The Brier skill score of this model is 0.088, representing the degree that this model improves over a naïve model (higher is better). The Brier reliability is 2.6E-4, representing distance to true probabilities (lower is better), while the resolution is 3.3E-3, representing forecast distances to the mean rate (higher is better). The SHAP plot shows that this model is most strongly driven by pre-procedural hemoglobin and coronary artery disease symptoms at presentation (**Figure 2**). The inclusion of both continuous variables and dichotomized show the importance of different values of this variable across decision trees in the final model.

### Decision 1: Access Site (Model 2)

The first decision point in this model is the choice of arterial access site: femoral or radial. When accounting for this decision, the AUROC improved to 0.817 and the AUPRC improved to 0.204. The Brier skill score improved to 0.091, the Brier reliability improved to 2.0E-4. The SHAP plot shows that femoral access is the ninth most informative variable in Model 2 (**Figure 3**). The inclusion of seemingly mutually exclusive variables (higher hemoglobin vs. lower hemoglobin) both being important is a function of different decision trees and described further in the **Supplementary Material**. Procedures performed via femoral access had a slightly increased rate of bleeding, while those performed via a radial access site had a variably decreased rate of bleeding. The fact that these procedures have similar SHAP values indicates that the model assigns similar risk to procedures with femoral access.

**Figure 3.**
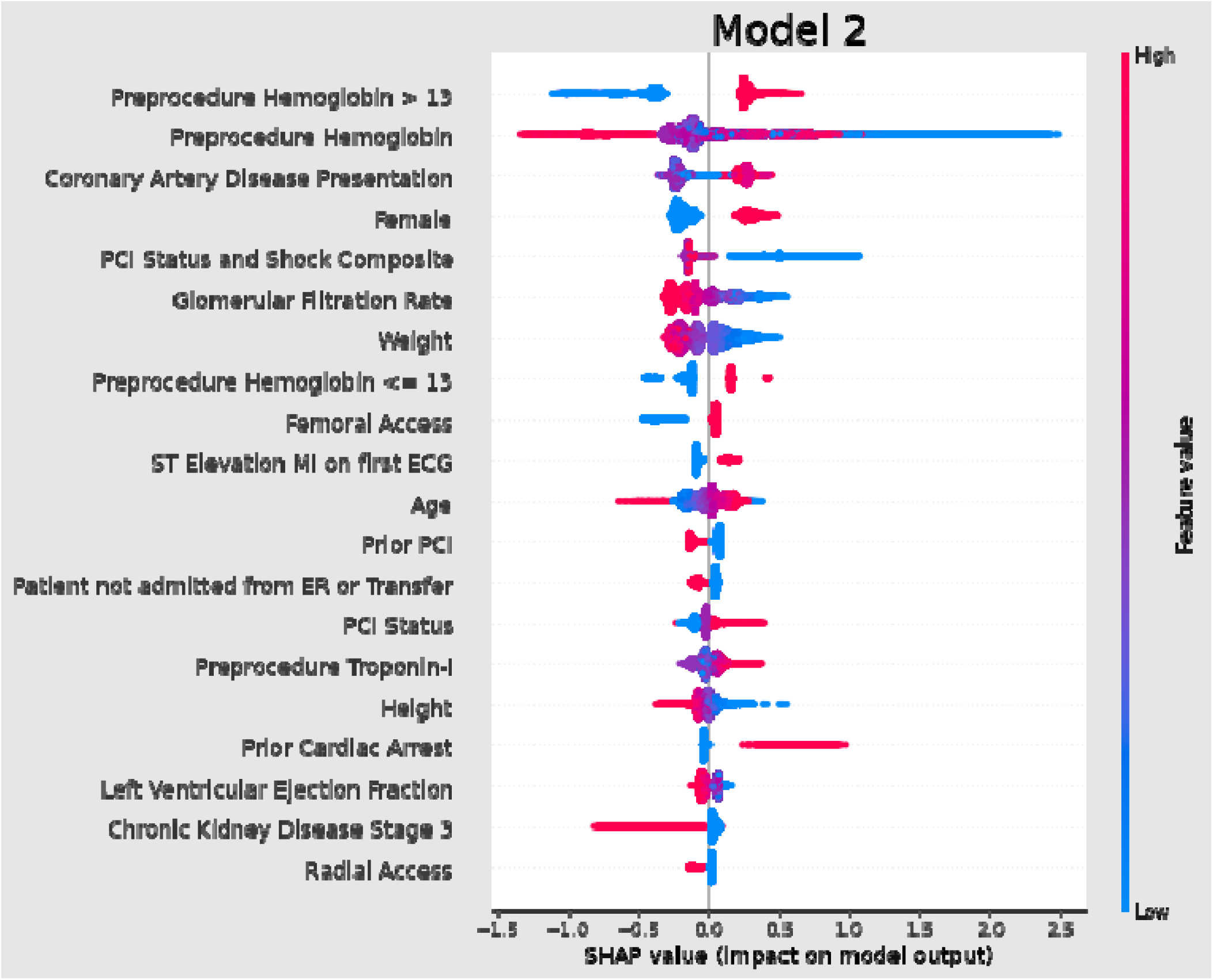
SHAP Tree explainer for Model 2. Procedures performed via femoral access are represented by the narrow red line to the right of the axis. In contrast, the blue points to the left of the axis represent procedures performed with radial access, and their elongated shape indicates that the radial access has a variable effect on bleeding risk, with the risk for some procedures being decreased by much more than the risk for others.

**Table 3A** presents a shift table describing patient risk categories(19). Among 123,712 patients classified as low (<1%) risk of bleeding by the clinical presentation model, 9,071 (7.3%) were reclassified as medium (1-4%) risk of bleeding by the model incorporating access site (**Table 3A**). Among those reclassified, 0.99% experienced a bleeding event. Among 270,485 patients classified as medium risk of bleeding by the initial model, 33,129 (12.2%) were reclassified as low risk by the subsequent model, while 6,465 (2.4%) were reclassified as high (>4%) risk. Of the 33,129 patients reclassified as low risk, 0.5% experienced a bleeding event, while of the 6,465 patients reclassified as high risk, 3.1% exhibited a bleeding event. Among 160,165 patients classified as high risk of bleeding by the initial model, 14,582 (9.1%) were reclassified as medium risk. Among those patients, 2.5% experienced a bleeding event.

**Table 3.** Shift tables following each decision point. Top value in each cell is number of patients classified into that risk bin by the two respective models. The bottom value in each cell indicates the actual bleeding rate of all patients within that cell. **Table 3** shows shift tables before and after each decision, while **Table 4** shows a shift table from the initial to final model. The earlier model is displayed left to right, while the later model is displayed top to bottom. The top number in each cell represents the number of patients assigned to that risk bin by each model. The bottom number in each cell is the overall bleeding rate of all patients in that cell. NaN represents that no patients were in that combination of bins.

| Initial vs After Access Site Decision |  |
| --- | --- |
|  | Model 1 |

|  | <1% | 1-4% | >4% | All |
| --- | --- | --- | --- | --- |
| **Model 2** | **Patients,**<br>**N**<br>**Observed**<br>**Rate** | **Patients,**<br>**N**<br>**Observed**<br>**Rate** | **Patients,**<br>**N**<br>**Observed**<br>**Rate** | **Patients,**<br>**N**<br>**Observed**<br>**Rate** |
| <1% | 114,641<br>0.57% | 33,129<br>0.54% | NaN | 147,770<br>0.56% |
| 1-4% | 9,071<br>0.99% | 230,891<br>1.69% | 14,582<br>2.54% | 254,544<br>1.72% |
| >4% | NaN | 6,465<br>3.11% | 145,583<br>10.14% | 152,048<br>9.84% |
| All | 123,712<br>0.60% | 270,485<br>1.58% | 160,165<br>9.45% | 554,362<br>3.64% |
| **In Cath Lab Before vs After Medication Decisions** |  |  |  |  |
|  | **Model 3** |  |  |  |
|  | <1% | 1-4% | >4% | All |
| **Model 4** | **Patients,**<br>**N**<br>**Observed**<br>**Rate** | **Patients,**<br>**N**<br>**Observed**<br>**Rate** | **Patients,**<br>**N**<br>**Observed**<br>**Rate** | **Patients,**<br>**N**<br>**Observed**<br>**Rate** |
| <1% | 140,103<br>0.43% | 32,764<br>0.64% | NaN | 172,867<br>0.47% |
| 1-4% | 11,244<br>1.20% | 207,541<br>1.67% | 21,612<br>3.24% | 240,397<br>1.79% |
| >4% | NaN | 11,448<br>4.37% | 129,650<br>11.22% | 141,098<br>10.67% |
| All | 151,347<br>0.49% | 251,753<br>1.66% | 151,262<br>10.08% | 554,362<br>3.64% |
| **Post PCI Before vs After Closure Decision** |  |  |  |  |
|  | **Model 5** |  |  |  |
|  | <1% | 1-4% | >4% | All |
| **Model 6** | **Patients,**<br>**N**<br>**Observed**<br>**Rate** | **Patients,**<br>**N**<br>**Observed**<br>**Rate** | **Patients,**<br>**N**<br>**Observed**<br>**Rate** | **Patients,**<br>**N**<br>**Observed**<br>**Rate** |
| <1% | 178,911<br>0.43% | 12,095<br>0.54% | NaN | 191,006<br>0.44% |
| 1-4% | 8,561<br>0.81% | 213,367<br>1.73% | 7,268<br>3.56% | 229,196<br>1.76% |
| >4% | NaN | 5,703<br>3.30% | 128,457<br>11.76% | 134,160<br>11.40% |
| All | 187,472<br>0.45% | 231,165<br>1.71% | 135,725<br>11.32% | 554,362<br>3.64% |

### Stage 2: Cardiac Catheterization Laboratory (Model 3)

The cardiac catheterization laboratory model uses information available after performing a diagnostic cardiac catheterization, but prior to initiation of PCI and choice of peri-procedural medications. In this model, bleeding prediction improved with an AUROC of 0.825 and an AUPRC of 0.208. The Brier skill score improved to 0.094, the Brier reliability worsened to 2.1E-4, and the Brier resolution improved to 3.5E-3. Of features added in this model, the SHAP plot shows that predictions are highly influenced by the presence of thrombus in a coronary lesion (13^th^ most informative variable) and pre-procedure Thrombolysis in Myocardial Infarction (TIMI) flow (14^th^ most informative variable) (**Figure 4**).

**Figure 4.**
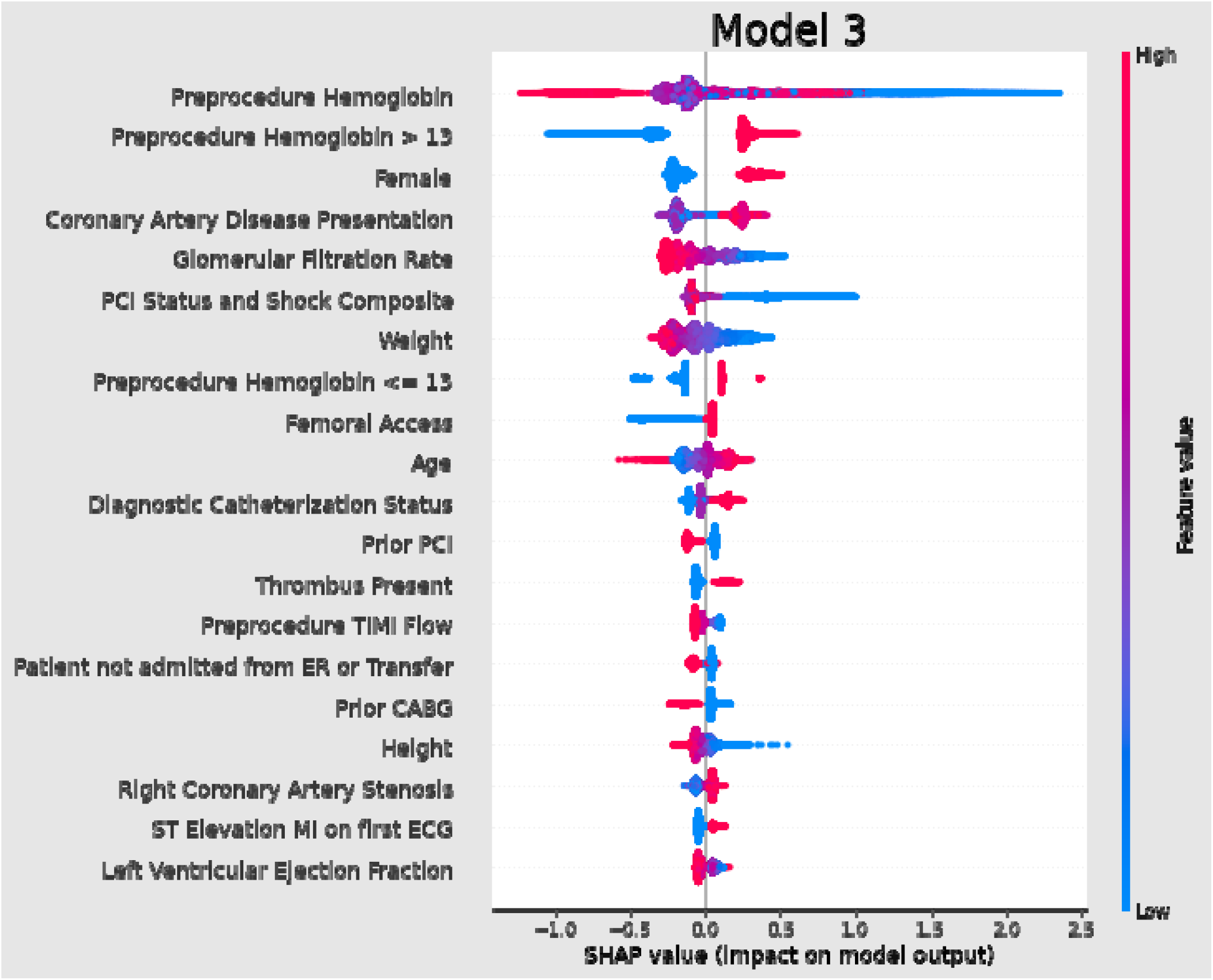
SHAP Tree explainer for Model 3.

### Decision 2: Intra-Procedure Medication (Model 4)

The next decision point is the choice of intra-procedural antiplatelet and anticoagulant agents. Following inclusion of these variables, the model performance increases to an AUROC of 0.832 and an AUPRC of 0.217. The Brier skill score improved to 0.102, the Brier reliability improved to 1.4E-4, and the Brier resolution improved to 3.7E-3. Of features added in this model the SHAP plot shows that use of glycoprotein IIb/IIIa inhibitors was strongly associated with an increased risk of bleeding (4^th^ most informative variable), while unfractionated heparin was less strongly associated with an increased risk of bleeding (14^th^ most informative variable) (Figure 5).

**Figure 5.**
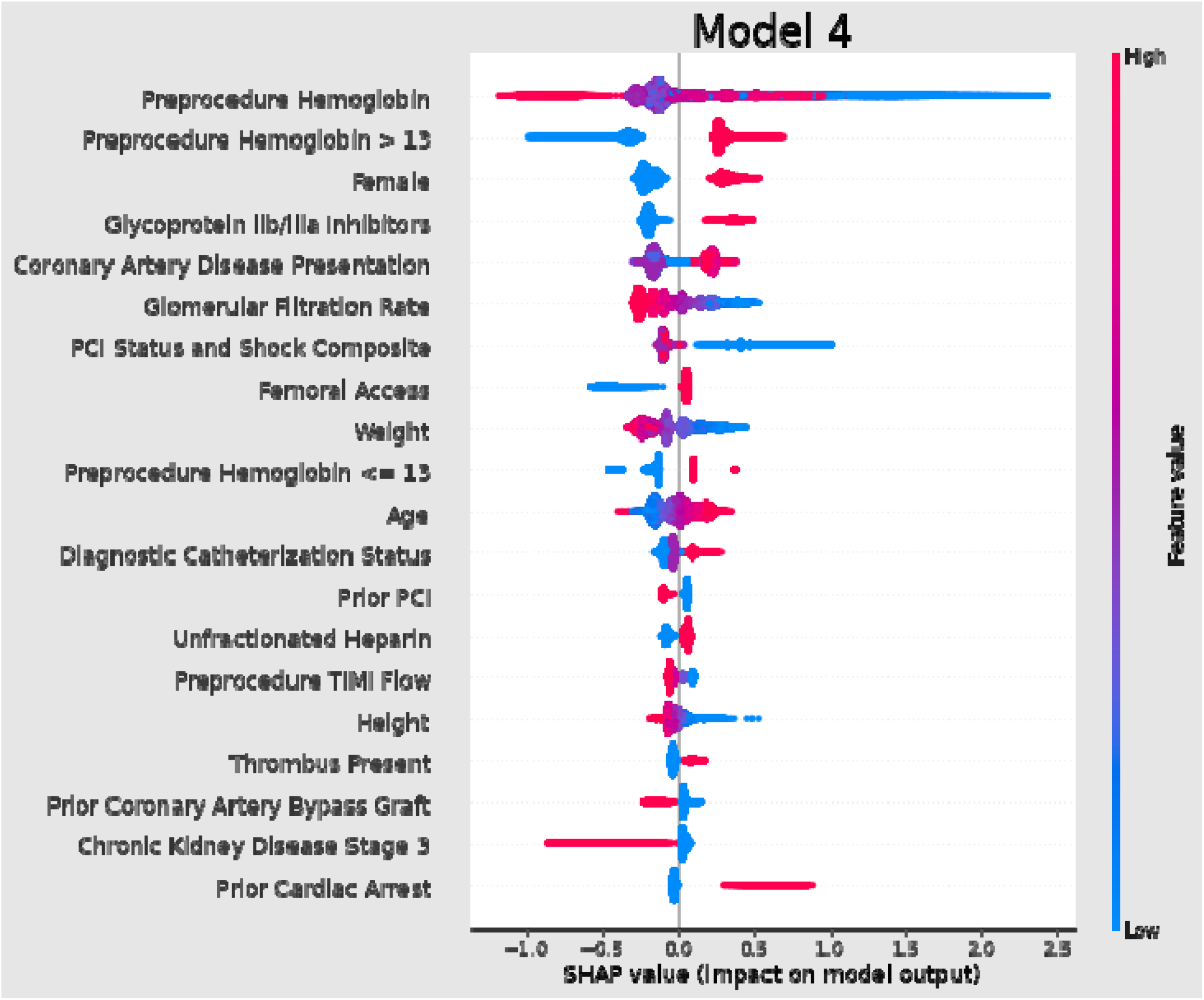
SHAP Tree explainer for Model 4

Among 151,347 patients classified as low risk of bleeding by the cardiac catheterization lab model (Model 3), 11,244 (7.6%) were reclassified as moderate risk by the model incorporating medication choices (Model 4), (Table 3B). Among those 11,244 reclassified patients, 1.2% experienced bleeding events, reflecting that these reclassifications improved model calibration. Among 251,753 patients classified as moderate risk of bleeding by Model 3, 32,764 (13.0%) were reclassified as low risk by Model 4, while 11,448 (4.5%) were reclassified as high risk. Among the 32,764 patients reclassified as low risk 0.6% experienced bleeding events, among the 11,448 patients reclassified as high risk 4.4% experienced bleeding events, also reflecting that the reclassifications improved calibration. Among 151,262 patients classified as high risk of bleeding by Model 3, there were 21,612 (14.3%) patients who were reclassified as moderate risk by Model 4. Among those patients, 3.2% experienced a bleeding event.

### Stage 3: PCI (Model 5)

The post-PCI model uses all information through PCI but prior to choice of closure method. This model improves upon the performance of prior models, with an AUROC of 0.844 and AUPRC of 0.241. The Brier skill score improved to 0.118, the Brier reliability improved to 1.1E-4, and the Brier resolution improved to 4.2E-3. The new features introduced in this model that were most associated with increased bleeding risk are proxies of PCI complexity and duration, including fluoroscopy time (10^th^ most informative variable) and contrast volume (20^th^ most important variable) (Figure 6).

**Figure 6.**
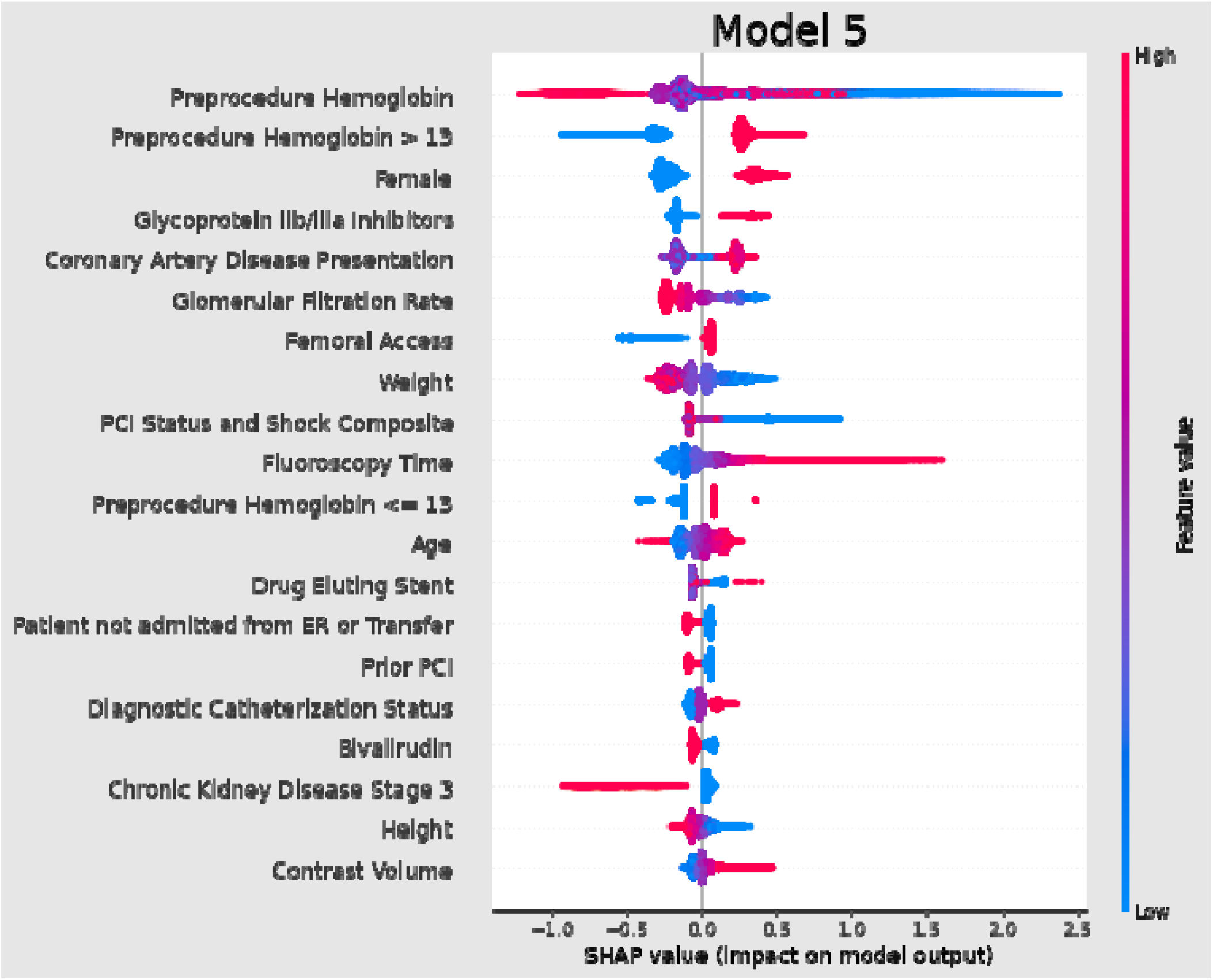
SHAP Tree explainer for Model 5

### Decision 3: Closure Method (Model 6)

The final decision point is closure. This decision had minimal effect on overall prediction, with AUROC remaining at 0.845 and AUPRC improving slightly to 0.242. The Brier skill score improved slightly to 0.119, the Brier reliability improved to 1.0E-4, and the Brier resolution improved to 4.3E-3. Figure 7 shows a SHAP explanatory plot for this model. Manual compression is the closure method most strongly predictive of increased bleeding risk (12^th^ most informative variable).

**Figure 7.**
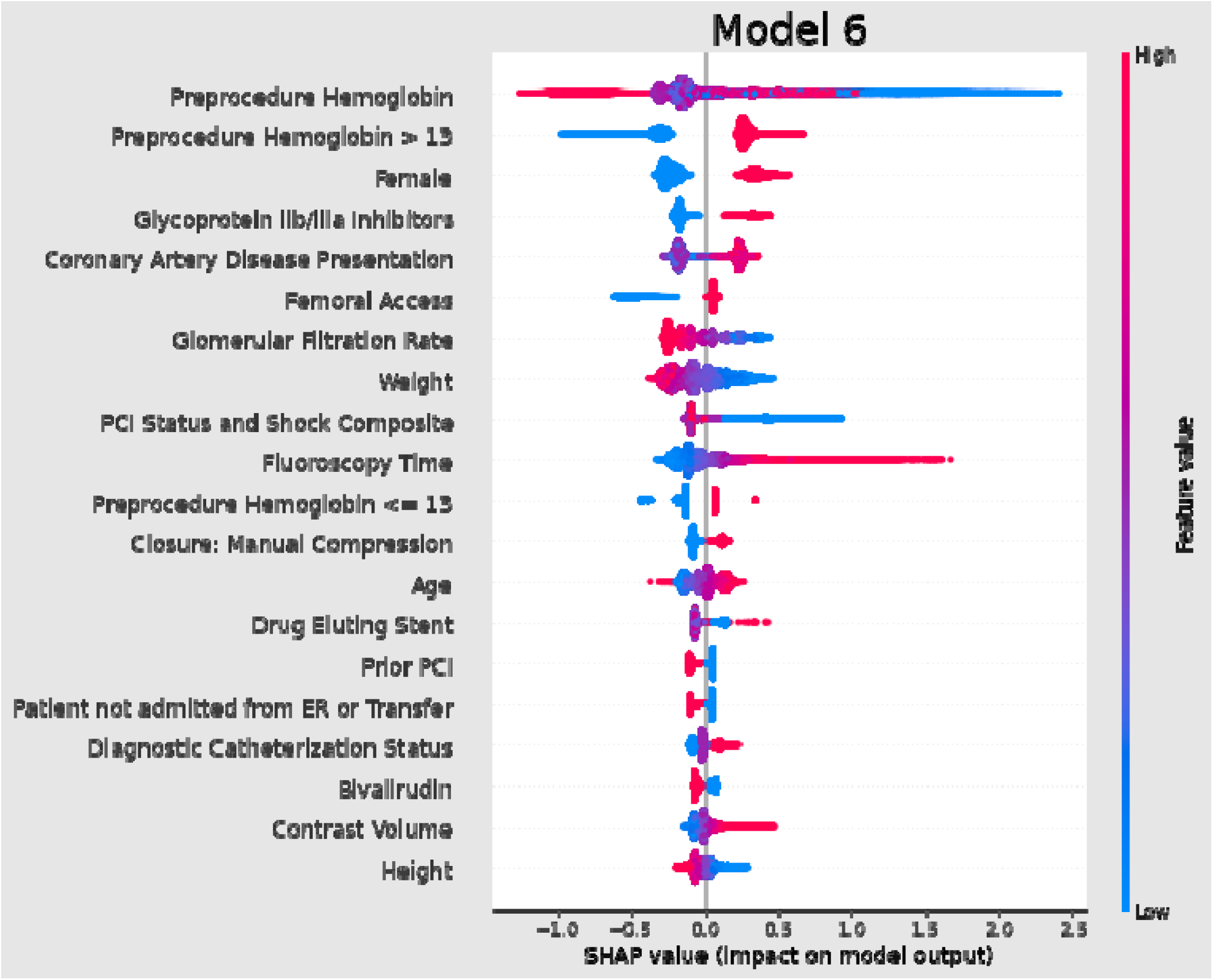
SHAP Tree explainer for Model 6

Among 187,472 patients classified as low risk of bleeding by the post-PCI model (Model 5), 8,561 (4.6%) were reclassified as moderate risk by the model incorporating closure decision (Model 6) (Table 3C). Bleeding events occurred in 0.8% of those patients. Among 231,165 patients classified as moderate risk of bleeding by Model 5, 12,095 (5.2%) were reclassified as low risk by Model 6, while 5,703 (2.5%) were reclassified as high risk. While the patients reclassified as low risk had an appropriately low occurrence of bleeding (0.54%), those patients reclassified to high risk had a moderate aggregated occurrence of bleeding (3.30%). Among 135,725 patients classified as high risk of bleeding by Model 5, there were 7,268 (5.4%) patients who were reclassified as moderate risk by Model 6. Bleeding events occurred in 3.6% of the reclassified patients.

### Reclassification from Model 1 to Model 6

Total reclassification from the initial model to the final model is shown in **Table 4**. Among 123,712 patients classified as low risk by the initial model, 14,441 (11.7%) were reclassified as moderate risk, while 723 (0.6%) patients were reclassified as high risk. Among the 14,441 patients re-classified as moderate risk, 1.4% experienced bleeding events. Among those 723 patients reclassified to high risk, 12.5% experienced bleeding events. Among 270,485 patients classified as moderate risk by the initial model, 82,418 (30.5%) were reclassified to low risk by the final model, while 16,577 (6.1%) were reclassified to high risk by the final model. Among the 82,418 patients reclassified to low risk 0.5% experienced bleeding events, while among the 16,577 patients reclassified to high risk 7.0% experienced bleeding events. Finally, among 160,165 patients classified as high risk by the initial model, there were 40 (<0.1%) patients reclassified to low risk, and 43,265 (27.0%) patients reclassified to moderate risk. The 40 patients reclassified to low risk experienced no bleeds (bleeding rate of 0%), while 2.5% of the 43,265 patients reclassified to moderate risk had a bleeding event.

**Table 4.**
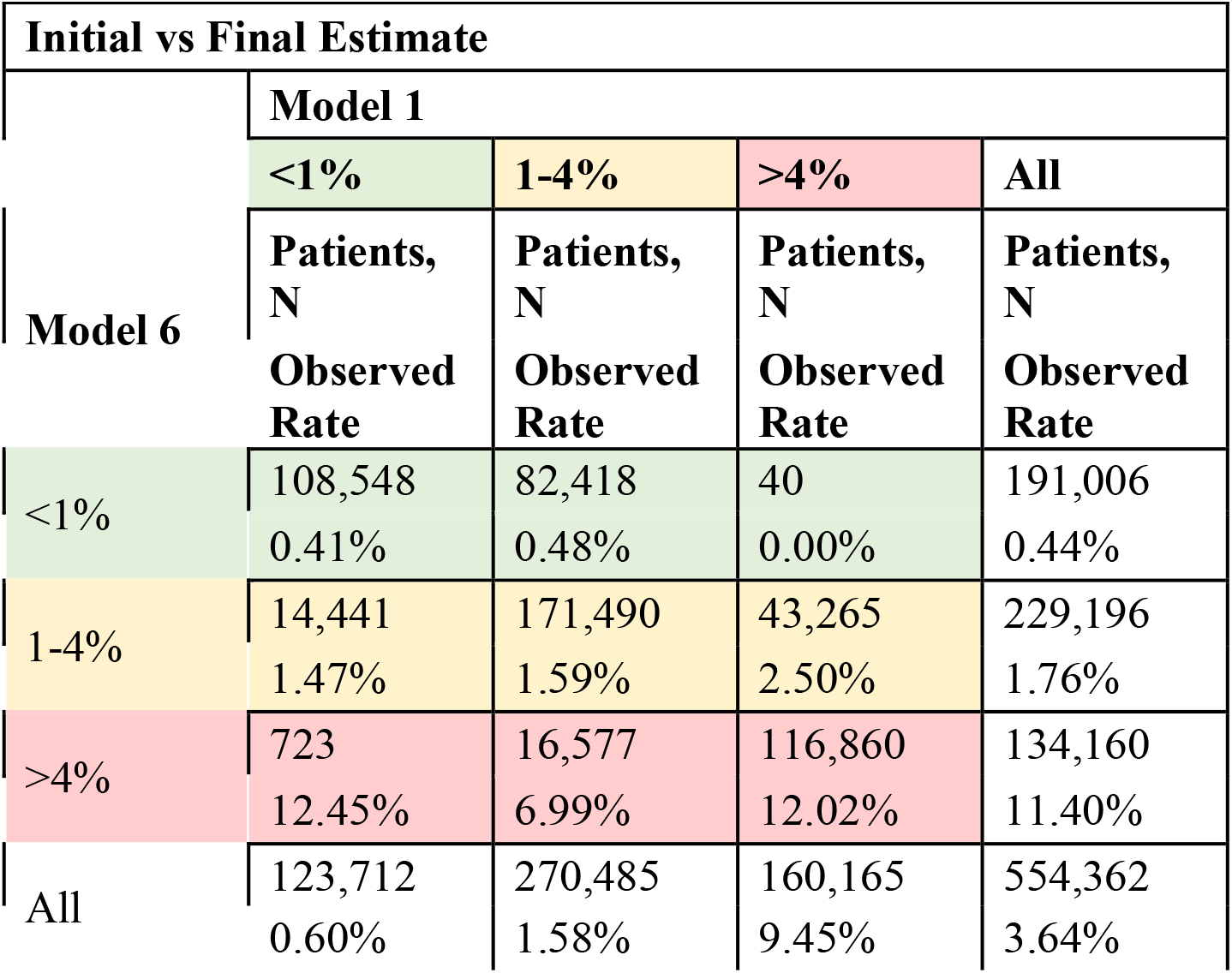
Shift table across models. Top value in each cell is number of patients classified into that risk bin by the two respective models. The bottom value in each cell indicates the actual bleeding rate of all patients within that cell.

## Discussion

In a large national registry, we demonstrate that the risk of post-PCI bleeding is dynamic and changes with treatment decisions. As additional data are gathered over time, estimates of this risk are meaningfully updated. We also found that as more data becomes progressively available, the model better identifies associations between variables and subsequent bleeding. The staged nature better represents an individual’s risk throughout the course of treatment and can inform treatment decisions in a way that is superior to using a pre-PCI model in isolation. As the model obtains more data, the association of this and the risk of bleeding increases in importance, as the decision itself represents a modest increase in risk at Model 2, but a larger increase in risk when additional data representing the case becomes apparent.

By extending beyond prior static machine learning research, which demonstrated that the full dynamic range of available variables allows higher-order, non-linear models to improve estimates of risk (4), this work demonstrates how the association of these variables in in all relevant formats (continuous and dichotomous) change in representation of risk throughout the course of care. Bleeding avoidance strategies were less used in those at highest risk based upon static models (25). While patients would receive only therapies with lower risk of bleeding, in some cases this must be balanced by antiplatelet and anticoagulant strategies that may reduce risk of coronary ischemia but with potential bleeding risk. Decision support that balances these factors may help to, therefore, inform other clinical decisions such as the PCI procedure itself (e.g., extent of revascularization performed may be more complete if a patient is at lower bleeding risk during the study).

To date, studies have failed to leverage the large numbers of variables tracked over an episode of care (8). Prior efforts to update models that estimate the risk of bleeding via machine learning still restricted utility of the model to a single decision point in time, using only the available data for when that decision would be made (4). This work identifies the dynamic nature of this bleeding risk and emphasizes the need for learning representations of risk over time. Additionally, this work identifies the utility of variables that may not have been appreciated to have an association with bleeding. The findings suggest the potential for additional improvement through direct integration with the electronic health record. By accessing data in near real-time, models will be able to present individualized estimates of risk and evaluation of treatment decisions, personalizing decision making and care throughout hospitalization and PCI.

### Limitations and Future Directions

There are several key limitations and opportunities for future research in our study. First, real-time data, which would be necessary for proactive implementation of dynamic models, can be challenging to acquire. The NCDR relies upon manual chart abstraction for many variables, such as past medical history variables. The implementation of such a system at scale within an electronic health record environment, where data may be available within near real-time capacity, requires additional investigation of natural language processing techniques, models that handle advanced time-series data, and appropriate evaluation of user interface design for reducing clinician burden when interacting with such a model.

The second is the timing of the available variables. The registry abstracts some variables as pre- and intra-procedure, so some confounding may exist from reactions to bleeds during the procedure. This nature of the variables could prevent accounting for some intra-procedural variables, such as if there was initially acute stent closure after PCI, which could require additional steps in the procedure.

Finally, the definition of the bleeding outcome includes multiple items in a composite form, which includes a hemoglobin cutoff. This suggests that patients with a low baseline hemoglobin were more likely to experience this outcome, because a small drop would put them below the threshold as opposed to a substantial drop being needed for other patients.

## Conclusion

We have developed a model that provides dynamically updated bleeding risk assessments by incorporating information available at different stages of patient care among patients undergoing PCI. These methods demonstrate evolution in variable importance as clinical decisions are made through course of care. For the risk of in-hospital bleeding, variable importance and bleeding risk changes as variables are included from cardiac catheterization and PCI. Accounting for the time-varying nature of data and capturing the association between treatment decisions and changes in risk provide up-to-date information that may guide individualized care throughout a hospitalization.

## Data Availability

All data produced in the present work are made available, by request, by the American College of Cardiology

https://cvquality.acc.org/NCDR-Home

## Abbreviations

PCI: percutaneous coronary intervention
XGBoost: eXtreme Gradient Boosting
AUROC: area under the curve of the receiver operating characteristic curve
AUPRC: area under the curve of the precision recall curve

## Perspectives

### Competency in Medical Knowledge

Understanding that risk of bleeding is dynamic, anti-coagulation decisions should constantly be re-evaluated through episodes of care.

### Translational Outlook 1

While static risk models may identify key driving risk factors, the dynamic nature of risk can inform decision making and provide updates based upon decisions made throughout course of treatment.

### Translational Outlook 2

Model interpretation techniques can better describe dynamic nature of risk.

## Acknowledgements

The views expressed herein represent those of the authors and do not necessarily represent the official views of the National Cardiovascular Data Registry or its associated professional societies identified online (https://cvquality.acc.org/NCDR-Home).

