## Supplemental Methods for "A dynamic model to estimate evolving risk of major bleeding after percutaneous coronary intervention"

**Supplementary material for “a dynamic model to estimate evolving risk of major bleeding after percutaneous coronary intervention”.**

**Hurley et al.**

**Supplementary Methods**

*Variables of Interest*

The current full existing NCDR bleeding risk model (3) uses 31 variables: 23 patient characteristics at the time of presentation and 8 characteristics related to coronary anatomy and lesion characterization. These variables included:

1. Two- or Three-vessel disease
2. STEMI
3. SCAI class II or III
4. SCAI class IV
5. Preprocedural TIMI flow grade is 0.
6. Left Main PCI
7. Prox LAD PCI
8. Subacute stent thrombosis

*Closure Devices and Radial Access*

The choice of closure device is a key indicator variable. While the choice of closure for radial access would be expected to be none, there are closure devices used with Radial Access. Without an ability to do a chart review, we take this data as is. 88% of Radial patients are coded as having a mechanical closure, 8% are coded as having manual compression and 4% as having a patch. The ability to verify the quality of the closure device data is a chief limitation of the staged-approach.

*Cleaning of data to correct missing data interpretation*

First, situations exist where a parent variable value of “no” indicates that daughter variables would not be captured (e.g., in a non-diabetic patient, no diabetic therapy is coded). Most daughter variables already had a category of missing, unknown, or other. We re-categorized the daughter variables to have a value of No/Not measured, and integrated “missing” for the few cases where the parent variable was a Yes/Measured variable and daughter variable was in fact missing.

Second, medications were categorized as no, yes, blinded (ie as a part of a clinical trial), or contraindicated. We re-categorized blinded as missing and re-categorized contraindicated as no.

Third, missing values were imputed using multiple multivariate feature imputation. Each missing feature was modeled using Bayesian ridge regressors trained in a round-robin fashion. Following imputation, binary and ordinal variables were set to the nearest allowed value. Multiple imputations were produced by sampling from the regressor models multiple times; each discrete sampling was a new overall sample from the model. This sampling was used to produce five folds of imputations.

*Why XGBoost*

XGBoost can evaluate higher order, non-linear interactions between variables automatically. This is necessary, since bleeding models based upon logistic regression selected the key variables based primarily on statistical tests between the variable and its relationship with incidence of major bleeding (3). While Logistic Regression can model with higher order, non-linear variables, these interaction terms need to be curated in advance.

*Precision and Recall Curves*

The area under the precision recall curve is directly impacted by the number of positive predictions made versus false positive or false negative estimates made, an important factor when considering cases with low event rates such as major bleeding (4.1%, **Table 1**).

**Supplementary Results**

*Why XGBoost Can Report Mutually Exclusive Variables as Important*

One might initially expect variables with the same concept to have only one side show key importance. For example, when hemoglobin is dichotomized, Logistic Regression would pick one and provide a coefficient value that describes the general model performance with respect to that risk factor. However, XGBoost, in the ability to make multiple decision trees to interpret importance across all types of cases, can have some trees where the higher hemoglobin variable is a stronger determining risk factor and in others, it might select the lower hemoglobin variable. While removing these to simplify interpretation may not impact model performance, they have been left as is to highlight the varied nature of risk across all participants, and for direct comparison with prior literature.

*Case Studies*

*Case Studies*

Because severe bleeding is a relatively rare event, risk changes for a minority of patients. However, some patients’ risk changes dramatically throughout the course of their care. Overall, the median difference of risk from the initial to final prediction is -0.41% (IQR -1.16%, +0.02%). However, the full range of risk changes was much larger, ranging from -44.4% to +83.2%. Two illustrative examples of more drastic changing risk is demonstrated in **Supplementary Figure 1** and described below.

Case Study A

A man in his 60s presented for emergent PCI for STEMI. In the initial model, his risk of bleeding was estimated to be 4.3%. This risk was driven predominantly by the emergent need for PCI, preprocedural hemoglobin, presence of STEMI, sex, and weight. The decision was made to use radial access, after which his bleeding risk was estimated to be 2.8%. There were no adverse findings during diagnostic coronary angiography, and risk decreased to 2.1%. He received prasugrel and unfractionated heparin, after which his risk fell to 1.5%. Following successful PCI, his risk further fell to 1.2%. His access site was closed using a suture-based closure device, and the final bleeding risk was 1.0%. This patient did not experience a bleed. Plots of individual SHAP values for this patient at each model stage are shown in **Supplementary Figure 2.**

Case Study B

A woman in her 60s presented for elective PCI for stable angina. In the initial model, her risk of bleeding was 0.9%. This risk was driven predominantly by her preprocedural hemoglobin, sex, stable nature of her coronary artery disease, and weight. Femoral access was used, after which her risk increased to 1.0%. Upon diagnostic coronary angiography, it was discovered that she had significant coronary stenosis present. The model at this stage estimated risk of bleeding to be 1.7%. She received unfractionated heparin and clopidogrel, after which her estimated risk was 1.4%. PCI was notable for high complexity (reflected by a long fluoroscopy time), after which her risk of bleeding increased to 5.1%. Her access site was sutured, and her final risk was 4.0%. This patient experienced a post-PCI bleed. Plots of individual SHAP values for this patient at each model stage are shown in **Supplementary Figure 3.**

**Discussion**

The changes in risk could inform optimizations in the approach to clinical decision-making prior to and during PCI. For example, the changes in model risk and importance in variables may provide for both confidence in decisions surrounding low- and high-risk participants and potential suggestions of additional data capture prior to making decisions for those in middle tiers of risk.

**Supplementary Figure 1.** Plot of case study risk scores across all model stages.

Case Study A began as high risk but was low risk in the final model. Case Study A did not ultimately bleed.

Case Study B began as low risk but was high risk in the final model. Case Study B ultimately experienced a bleed.
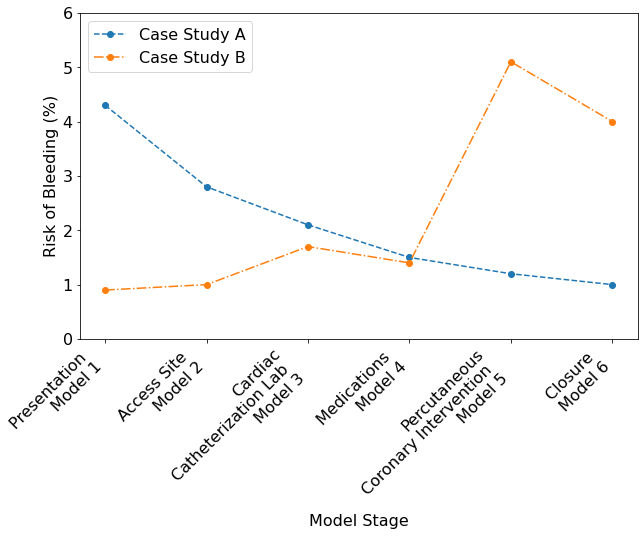

**Supplementary Figure 2.** SHAP explainer for Case Study A. At each model stage, the prediction is created by summing each variable contribution to risk. Variables on the left (red) are contribute to an increased risk of bleeding, while variables on the right (blue) contribute to a decreased risk of bleeding. Variables are organized such that those providing the strongest change to risk are at the center, with variables providing smaller changes to risk at the outside.
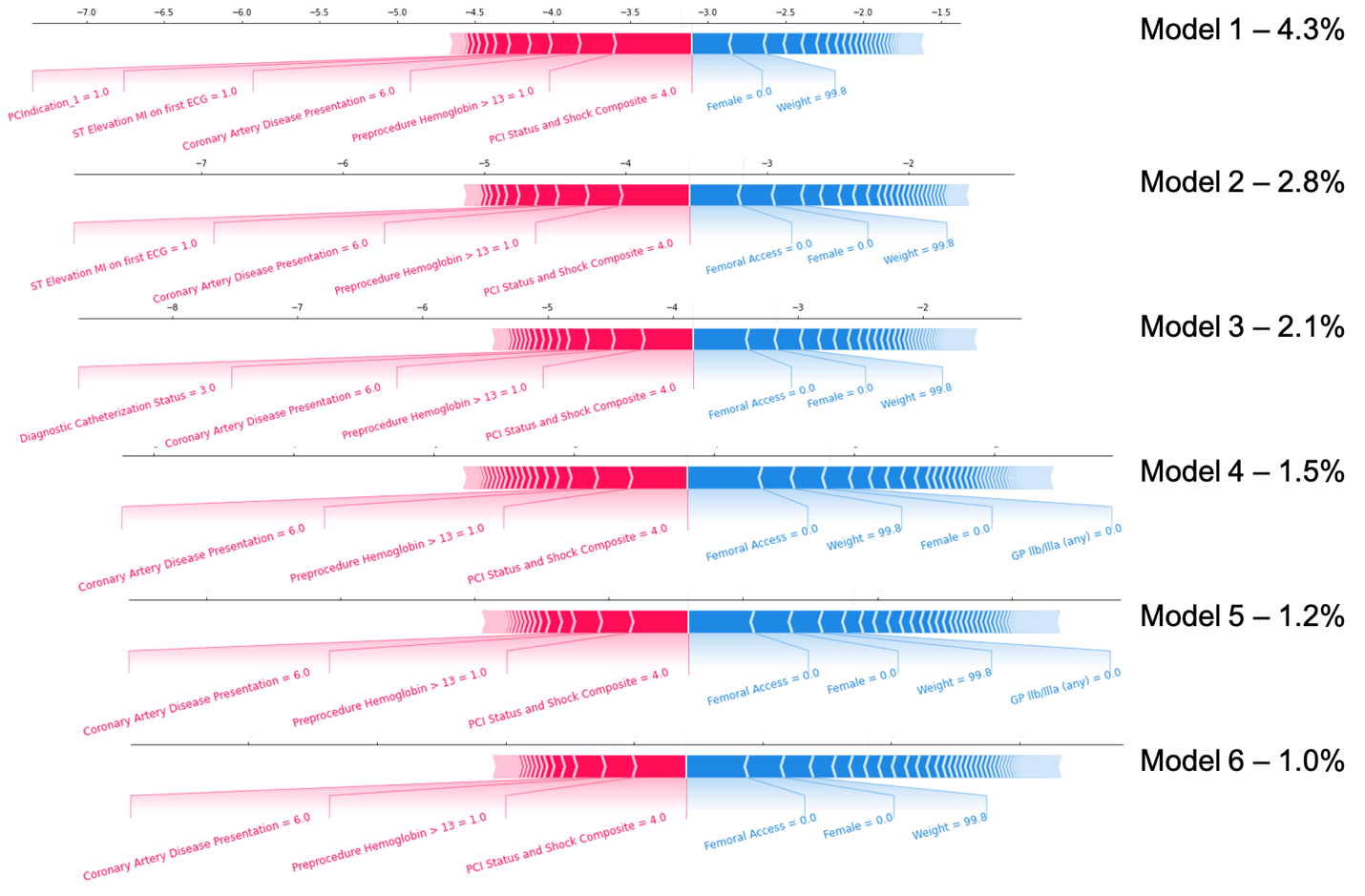

**Supplementary Figure 3.** SHAP explainer for Case Study B.
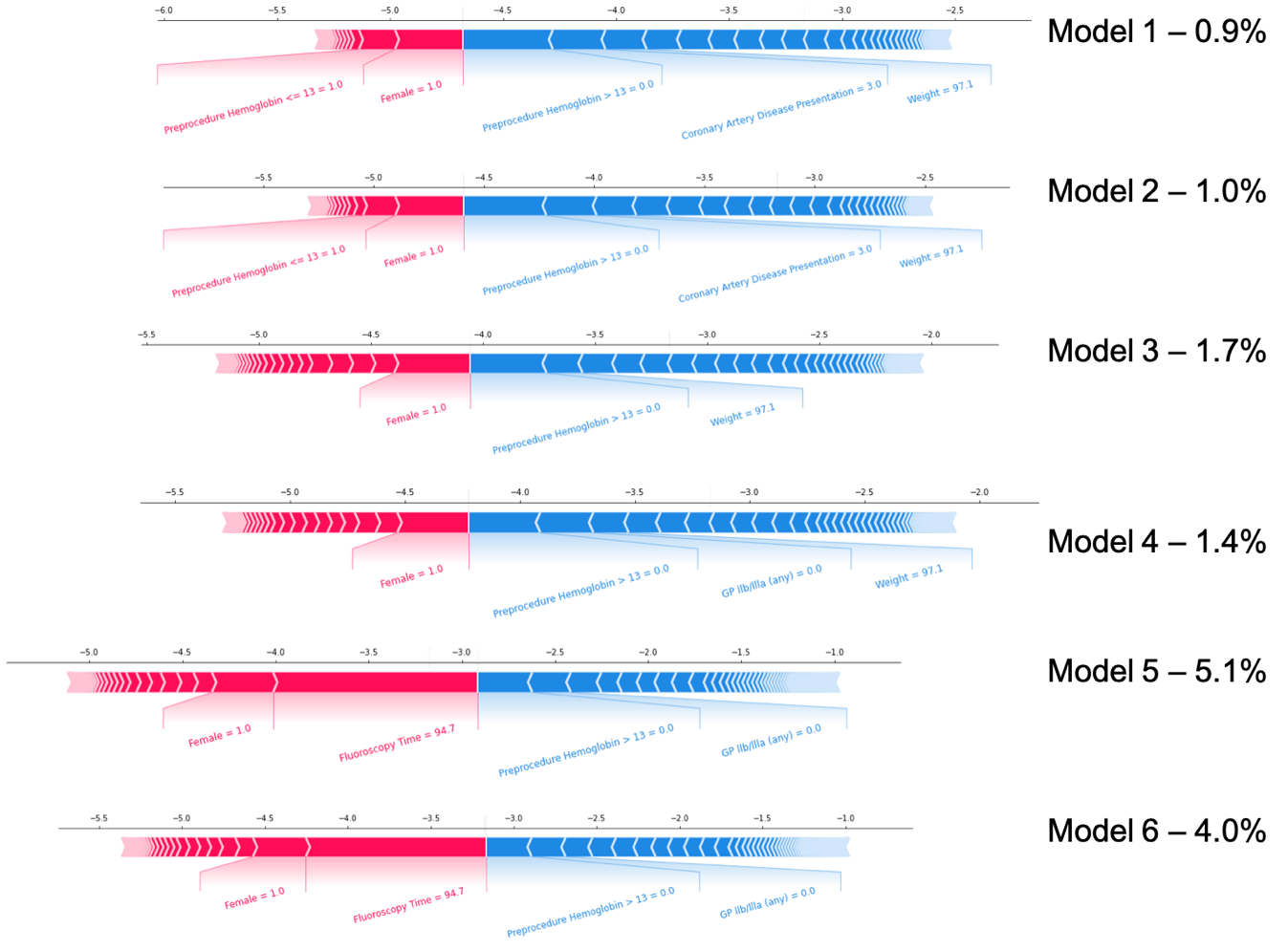
